# Quantifying bias of COVID-19 prevalence and severity estimates in Wuhan, China that depend on reported cases in international travelers

**DOI:** 10.1101/2020.02.13.20022707

**Authors:** Rene Niehus, Pablo M. De Salazar, Aimee R. Taylor, Marc Lipsitch

## Abstract

Risk of COVID-19 infection in Wuhan has been estimated using imported case counts of international travelers, often under the assumption that all cases in travelers are ascertained. Recent work indicates variation among countries in detection capacity for imported cases. Singapore has historically had very strong epidemiological surveillance and contact-tracing capacity and has shown in the COVID-19 epidemic evidence of a high sensitivity of case detection. We therefore used a Bayesian modeling approach to estimate the relative imported case detection capacity for other countries compared to that of Singapore.We estimate that the global ability to detect imported cases is 38% (95% HPDI 22% - 64%) of Singapore’s capacity. Equivalently, an estimate of 2.8 (95% HPDI 1.5 - 4.4) times the current number of imported cases, could have been detected, if all countries had had the same detection capacity as Singapore. Using the second component of the Global Health Security index to stratify likely case-detection capacities, we found that the ability to detect imported cases relative to Singapore among high surveillance locations is 40% (95% HPDI 22% - 67%), among intermediate surveillance locations it is 37% (95% HPDI 18% - 68%), and among low surveillance locations it is 11% (95% HPDI 0% - 42%). Using a simple mathematical model, we further find that treating all travelers as if they were residents (rather than accounting for the brief stay of some of these travelers in Wuhan) can modestly contribute to underestimation of prevalence as well. We conclude that estimates of case counts in Wuhan based on assumptions of perfect detection in travelers may be underestimated by several fold, and severity correspondingly overestimated by several fold. Undetected cases are likely in countries around the world, with greater risk in countries of low detection capacity and high connectivity to the epicenter of the outbreak.

## Introduction

During the outbreak of a new virus SARS-Cov2 and its associated disease COVID-19, infection in travelers has been used to estimate the risk of infection in Wuhan, Hubei Province, China, the epicenter of the outbreak^1^. This approach is similar to that used for the the 2009 influenza pandemic where infections in tourists returning from Mexico were used to estimate the time-specific risk of infection (incidence or cumulative incidence) with the novel pandemic H1N1 influenza strain in Mexico (or parts thereof). The idea was that surveillance for the novel virus was not intense during the early days of the pandemic in Mexico, the source country, and that detection would be far more sensitive in travelers leaving Mexico, who would be screened when returning home as a means of preventing introductions of cases into destination countries ^2,3^. Reports that health systems in Wuhan are overwhelmed and many cases are not being counted have led to the use of outgoing traveler data to estimate the time-specific risk of COVID-19 in Wuhan^4^. This estimate, in turn, has been used to estimate the cumulative incidence of infection by a certain date in Wuhan, and from there (often assuming exponential growth and no appreciable depletion of susceptibles) the cumulative number of cases. Two important assumption underlie this calculation: i) that the detection of cases in the destination country has been 100% sensitive and specific, whether they are detected at the airport (prevalent cases with symptoms) or later after arrival at their destination (cases that were incubating during travel); ii) that travelers have the same prevalence of infection as the average resident of Hubei, so the prevalence inferred in travelers may be directly applied in Hubei. Here we consider the extent to which these two assumptions are justified. We conclude that the first assumption is strongly inconsistent with observed data, resulting in potentially substantial underestimates of prevalence in Hubei and corresponding overestimates of case-severity measures that are normalised by case counts.

We previously showed that there was variability between locations in the world in the relationship between the number of travelers from Wuhan to each international destination and the number of imported cases detected in that destination. On average, for countries presumed to have high surveillance capacity, one imported case reported over the period 8th January to 4th February was associated with each additional 14 passengers/day historical travel volume^5^. However there was variation around this average. Among countries with substantial travel volume, Singapore showed the highest ratio of detected imported cases to daily travel volume, a ratio of one case per 5 daily travelers. Singapore is historically known for exceptionally sensitive detection of cases, for example in SARS^6^, and has had extremely detailed case reporting during the COVID-19 outbreak^7^. We therefore use Singapore in this work as the upper limit of case detection capacity. And we estimate this capacity of other locations relative to Singapore.

Regarding the second assumption, we demonstrate that the point prevalence of infection may be lower in visitors who have stayed only briefly in the source population (Wuhan) than in residents. All else equal, the discrepancy between resident and visitor prevalence is most pronounced if the visitors’ durations of stay are shorter, for slower-growing epidemics, and for longer durations of detectable infection; conversely, visitors are more similar to residents in their prevalence of infections if they stay longer, if the epidemic is growing faster, and if the duration of detectable infection is shorter. We quantify this discrepancy as a function of these features using a simple model.

## Methods

### Data

From a total of 195 worldwide locations (reflecting mainly countries without taking any position on territorial claims), we included *n*=194, which excludes the epicenter China. Data on imported cases aggregated by location were obtained from the WHO technical report dated 4th February 2020^1^ (a zero case count was assumed for all locations not listed). We used case counts up to the 4th February, because after this date the number of exported cases from Hubei province drops rapidly^1^, likely due to the Hubei-wide lockdowns. We defined imported cases as those with known travel history from China (of those, 83% had travel history from Hubei province, and 17% from unknown locations in China^1^). Estimates on daily air travel volume were obtained from Lai et al.^8^. They are based on historical (February 2018) data from the International Air Travel Association and include estimates for the 27 locations that are most connected to Wuhan. They capture the daily average number of passengers traveling via direct and indirect flight itineraries from Wuhan to destinations outside of China. For all 167 locations not listed by Lai et al.^8^, we set the daily air travel volume to 1.5 passengers per day, which is one half of the minimum reported by Lai et al. Surveillance capacity was assessed using the Global Health Security Index^9^, which is an assessment of health security across 195 countries agreeing to the International Health Regulations (IHR [2005]). Specifically, we use the second category of the index, Early Detection and Reporting Epidemics of Potential International Concern, henceforth referred to as simply the GHS_2_ index. We classify locations with GHS_2_ index above the 80th percentile as high surveillance locations, those with GHS_2_ index below the 20th percentile as low surveillance locations, with all others classified as locations with intermediate surveillance. We treat Singapore as a special case for surveillance of COVID-19, and we assign it it’s own category of highest-achieved surveillance. This study did not include human subjects, used publicly available data, and therefore no ethical approval was required.

### Estimating detection probability relative to Singapore

We consider the detection of 18 cases by 4th February 2020 in Singapore^1^ to reflect the highest surveillance capacity among all locations, and estimate the probability of detection in other countries relative to Singapore according to the following model. We model the case detection across *i* = 1,…,*n* worldwide locations, where the *n* = 194 locations are indexed with Singapore being *i* = 1, followed by the rest of the locations in order of decreasing GHS_2_ index. We assume that the observed case count across the *n* locations follows a Poisson distribution, and that the expected case count is linearly proportional to the daily air travel volume and a random variable, θ_*level*_, reflecting the capacity of detecting cases relative to Singapore:

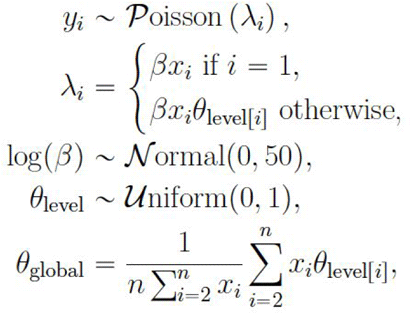

where *y*_*i*_ denotes the reported case count in the *i*-th location, *λ*_*i*_ denotes the expected case count in the *i*-th location, *β* denotes the regression coefficient, *x*_*i*_denotes the daily air travel volume of the *i*-th location, and θ_*level*_ denotes a proportion of Singapore’s detection capacity, with θ_*level*[*i*]_ being a realisation of θ_*level*_ for location *i*. We assume that there are three different levels: low, medium and high. For each θ_*level*_ ∈ {θ_*low*_, θ_*med*_, θ_*high*_} we assign a uniform prior over [0,1], and for log(*β*) we assign a weakly informative Normal prior with mean zero and standard deviation 50. Having fit the model (see details below) we approximate the distribution of the average detection probability, realisations of which, θ_*global*_, are obtained by transforming draws from the posterior distributions of θ_*low*_, θ_*med*_, θ_*high*_. Specifically, we take the weighted mean of the posterior draws of θ_*low*_, θ_*med*_, θ_*high*_ for *i* = 2,…*n* where weights are proportional to daily air travel volume, *x*_*i*_. Exclusion of Singapore (*i*=1) enables the estimation of the global detection probability relative to Singapore. Conversely, 1/θ_*global*_ is an estimate of the multiplier of the case count that could have been detected globally under a capacity equivalent to that of Singapore. We discuss the mean and 95% highest posterior density interval (HPDI) of the numerical approximation of the posterior distribution of θ_*global*_, as well as the mean and 95% HPDI of the numerical approximation of the posterior distribution of 1/θ_*global*_. Note that the latter two are not reciprocals of the former two because the inverse of a mean is not equal to the mean of the inverse, and similarly for the HPDIs.

We fit this model using Stan software (v2.19.1)^10^ and we draw 80,000 samples from the posterior using four independent chains (20,000 samples each), each with a burn-in of 500. Diagnostic plots of the MCMC sampler for each of the inferred variables (θ_*low*_, θ_*med*_, θ_*high*_ and β) are shown in **Supplementary Figure 1**. All analyses are fully reproducible with the code available online (https://github.com/c2-d2/detect_prob_corona2019).

### Testing the effect of length of stay in point prevalence of travelers

In 2009, during the influenza pandemic which originated in Mexico, it was assumed that most travelers leaving Mexico were tourists, or other temporary visitors, with relatively short stays in Mexico, and that the risk that they were infected represented a cumulative hazard over the period of their stay^2,3^. The basic assumption was that short term visitors faced the same *hazard* of infection as residents of Mexico, but, given the shorter stay, they had a somewhat lower *prevalence* of infection when returning to their home country. Many estimates in 2019-20 for COVID19 have instead made the assumption of equal prevalence in travelers leaving Wuhan and in residents, which is equivalent to assuming either that all travellers are Wuhan residents, or that all visitors had stayed long enough during the epidemic that their prevalence was similar to that of residents.

To quantify the difference between these two scenarios – assuming that all travellers are short term visitors versus assuming that all travellers are residents or long term visitors – we considered a simple model of an exponentially growing epidemic, in which the hazard of infection at time *t* is *λ*(*t*) increasing exponentially at rate *r*. At the beginning of the epidemic, which we call time 0, the hazard of infection is *λ*(0) and thereafter *λ*(*t*) = *λ*(0)*e* ^*rt*^. Then the point prevalence of infection at time *u* in residents who have stayed in Wuhan for the duration of the epidemic will be the probability that they have become infected and not recovered by time *u*, assuming that the cumulative hazard remains small enough by that point that there has been no appreciable depletion of susceptibles:

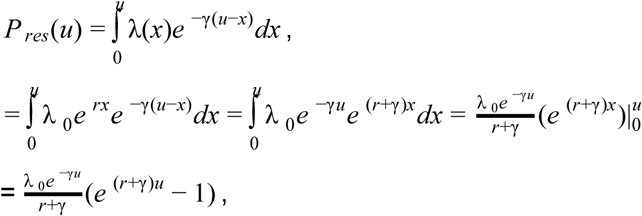

assuming exponentially distributed duration of detectable infectiousness, with mean duration γ ^−1^. The same quantity for a visitor who had only been in Wuhan for *d* days prior to departure would be 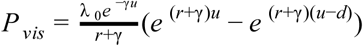 assuming that they differ from residents only in the duration of exposure, not in the intensity of exposure. Under these assumptions, the ratio of prevalence in visitors to that in residents, which we call *V*, would be 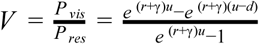. Once the number of cases is substantial, this term can be well approximated as *V* ≈ 1 − *e*^−(*r*+*γ*)*d*^ We plot this approximation of *V* given doubling times aligned with ^4^, a range of durations of detectable infection and a range of lengths of stay^2^ (**Figure 2**).

**Figure 1.**
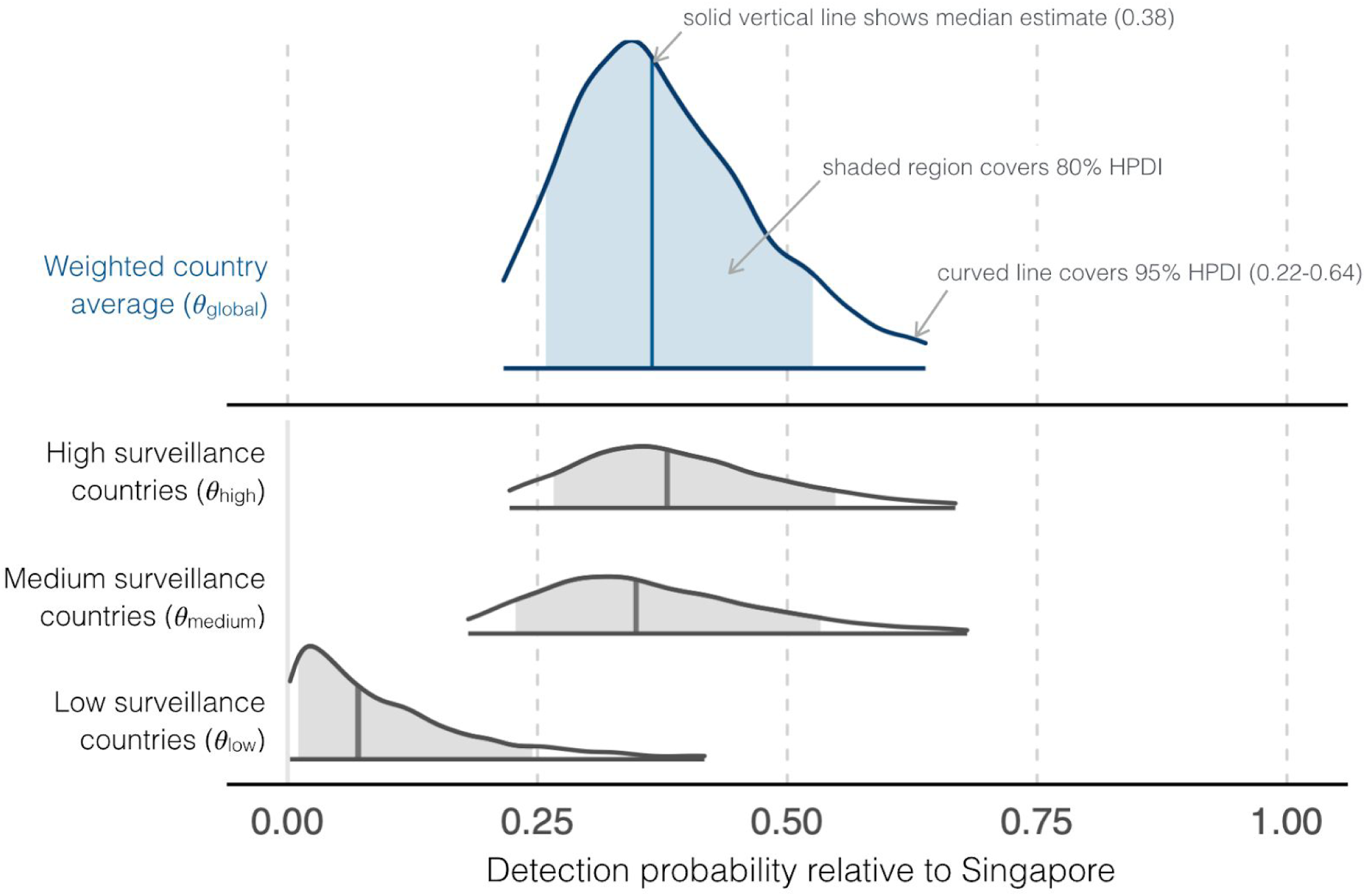
Posterior distributions of detection probabilities relative to Singapore. The bottom panel shows the posterior distributions of θ_*low*_, θ_*med*_, θ_*high*_. The top panel is a density plot of θ_*global*_. HPDI: Highest Posterior Density Interval.

**Figure 2.**
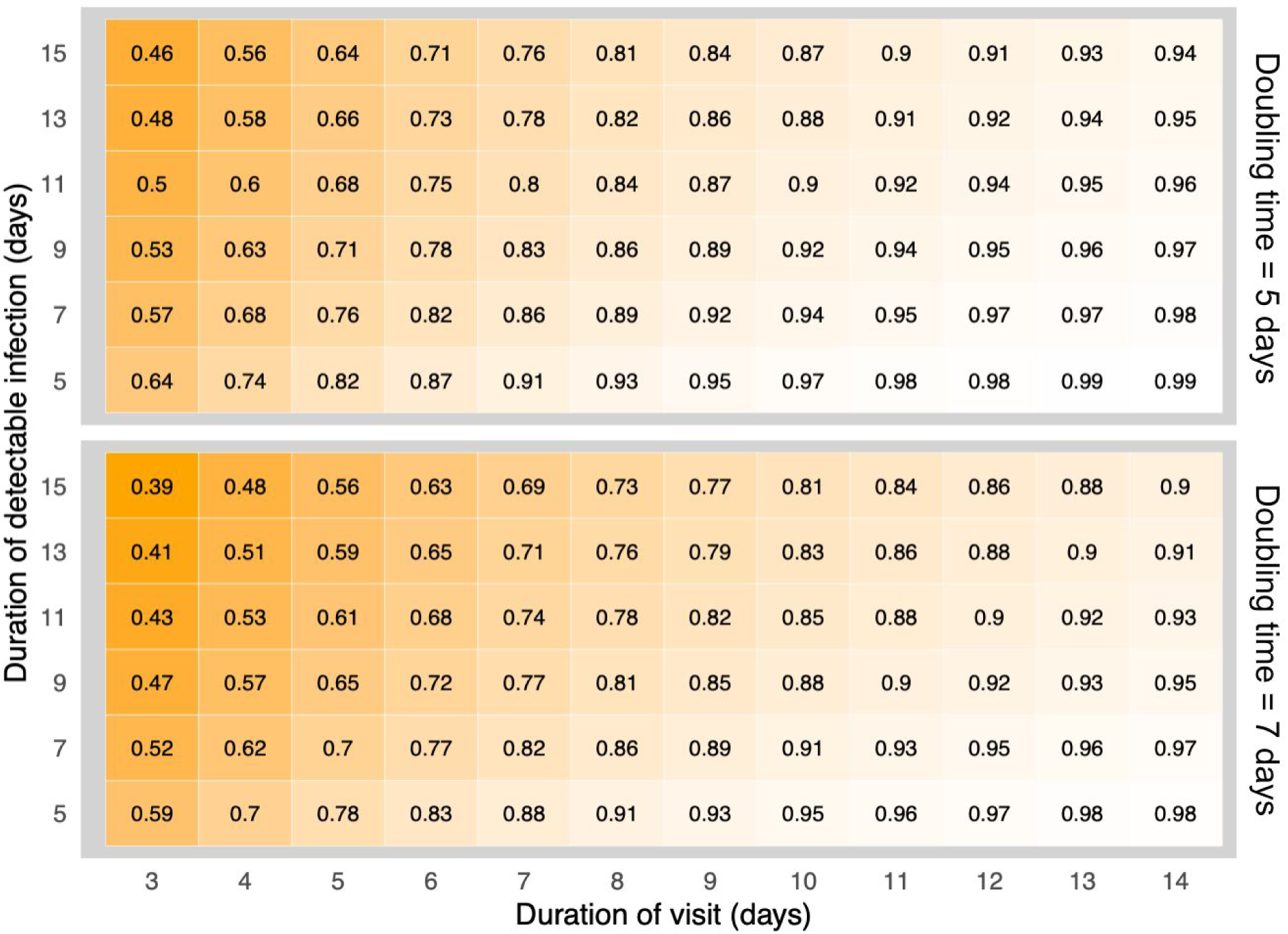
Ratio of infection prevalence in temporary visitors relative to that in residents (*V*). Plot shows *V* over a range of durations of visit (*d*) and a range of durations of detectable infection (γ ^−1^). In the upper panel the doubling time of the epidemic is 5 days and in the lower panel it is 7 days. The ratios are given as numbers (rounded to two decimals) with lighter areas as *V* approaches 1.

## Results

We estimate that the global ability to detect imported cases is 38% (95% HPDI 22% - 64%) of Singapore’s capacity. Equivalently, an estimate of 2.8 (95% HPDI 1.5 - 4.4) times the current number of imported cases, could have been detected, if all countries had had the same detection capacity as Singapore, which leads to 1.8 (95% CI 0.5 - 3.4) undetected cases per detected case. The ability to detect imported cases among high surveillance countries is 40% (95% HPDI 22% - 67%), among intermediate surveillance countries it is 37% (95% HPDI 18% - 68%), and among low surveillance countries it is 11% (95% HPDI 0% - 42%).

We further find that the prevalence ratio between temporary visitors and residents approaches 1 as the epidemic growth rate, the duration of stay, and the recovery rate increase and it approaches zero for short duration of stay, long duration of infection, and slower epidemic growth (**Figure 2**).

## Discussion

In this paper, we have aimed to test two assumptions underlying the estimation of incidence at the epicentre of the SARS-Cov2 outbreak. The first of these is that the capacity for detection of international imported cases is 100% sensitive and specific across locations. While we know of no reason to doubt specificity of detection, we tested the assumption of perfect sensitivity. Based on our previous estimates^5^ we assumed Singapore has the highest capacity of surveillance with respect to COVID-19. We regressed the cumulative cases against Wuhan-to-location air travel volume considering Singapore to have the greatest detection capacity and estimating the relative underdetection compared to Singapore in the remaining 190 locations classified according to the GHS_2_ index^9^. While it is unlikely that this index reflects the true ranking of countries for any specific epidemic, such as the current one of COVID-19, we assume that it captures different levels of surveillance capacity. We therefore grouped the remaining locations into three big classes high, medium, and low surveillance capacity instead of using exact ranking.

We estimated that detection of exported cases from Wuhan worldwide is 38% (95% HPDI 22%-64%) as sensitive as it has been in Singapore. Put another way, this implies that the true number of cases in travelers is at least 2.8 (95% HPDI 1.5 - 4.4) times the number that has been detected. Equivalently, for each detected case there are at least 1.8 (95% CI 0.5-3.4) undetected cases. If the model is correct, this is an upper bound on the detection frequency because (1) Singapore’s detection is probably not 100% efficient. Singapore had as of 12 February 2020 eight documented cases of COVID-19 transmission for which there were no known epidemiological links to China or other known cases^11^, implying that imported cases in Singapore may have gone undetected (although it is not certain that these imports came from Wuhan or China, and links may still be found). (2) Singapore’s detection like that in other countries has relied largely on symptoms and travel history, so the number of asymptomatic or low-severity cases missed by such a strategy is unknown.

The second assumption we tested is that the true prevalence in travelers is similar to that of residents. It may be different for either of two reasons, one of which we attempt to quantify here. It could be less if those who travel for some reason are less well integrated into the social mixing that produces infection, for example if they tend to have stayed in certain parts of the city or in hotels. This aspect could conceivably increase or decrease prevalence in travelers relative to residents. Here we quantify a second difference, which is that some travelers (whom we refer to as visitors) will have been in the city only for a short time and had less exposure to the infection than residents. This effect, we find, is most pronounced when the epidemic is growing slowly, when the visitors have stayed only briefly, and when the duration of detectable infection is short. We find that for plausible parameters for COVID-19, prevalence in visitors staying only 3 days could be as little as half that of residents, but for longer stays of over a week the visitor prevalence should be 80% or more that of residents. Assuming that the traveler population is a mix of visitors of various durations and residents, this suggests that underestimation of source population prevalence due to the presence of short-stay visitors could be appreciable but more modest than the effect of imperfect detection.

These findings that detected cases in travelers likely underrepresent the source population prevalence have two important implications for public health response to SARS-CoV2. First, this finding has implications for approaches to case burden and severity estimation which use cases in travelers to impute cases in Wuhan, which are then compared (for severity estimation) against deaths in Wuhan. If the true number of cases in travelers is higher than previously thought, this implies more cases in Wuhan and a larger denominator, resulting in reduced estimates of severity compared to estimates assuming perfect detection in travelers. Future studies should account for our evolving understanding of detection capacity when estimating case numbers and severity in source population on the basis of traveler case numbers. Second, the scenario where the virus has been imported from Wuhan and remained undetected in various worldwide locations is a plausible one, at least until the city lockdown (23rd January 2020), and one might speculate that detection capacity remained limited beyond this period as travelers infected elsewhere in China continued to leave China. Based on our model, the risk of undetected circulation correlates both with air travel connectivity and (inversely) to outbreak detection capacity, but could have happened in virtually any location worldwide leading to the potential risk of self-sustained transmission, which may be an early stage of a global pandemic.

## Data Availability

All analyses are fully reproducible with the code available online

https://github.com/c2-d2/detect_prob_corona2019

## Role of the funding source

The funding bodies of this study had no role in the study design, data analysis and interpretation, or writing of the manuscript. The corresponding authors had full access to all the data in the study and had final responsibility for the decision to submit for publication.

## Contributors

RN, PMS, and ML designed the study. All authors performed the analysis, reviewed and approved the final manuscript.

## Declaration of interests

Marc Lipsitch has received consulting fees from Merck. All other authors declare no competing interests.

## Acknowledgments

We thank Shengjie Lai for providing the air travel data, and we thank Nicholas Jewell and Stephen Kissler for valuable feedback along the way. This work was supported by Award Number U54GM088558 from the US National Institute Of General Medical Sciences. P.M.D was supported by the Fellowship Foundation Ramon Areces. A.R.T. was supported by a NIGMS Maximizing Investigator’s Research Award (MIRA) R35GM124715-02. The content is solely the responsibility of the authors and does not necessarily represent the official views of the National Institute Of General Medical Sciences or the National Institutes of Health.

## Appendix

**Supplementary Figures**

**Supplementary Figure 1.**
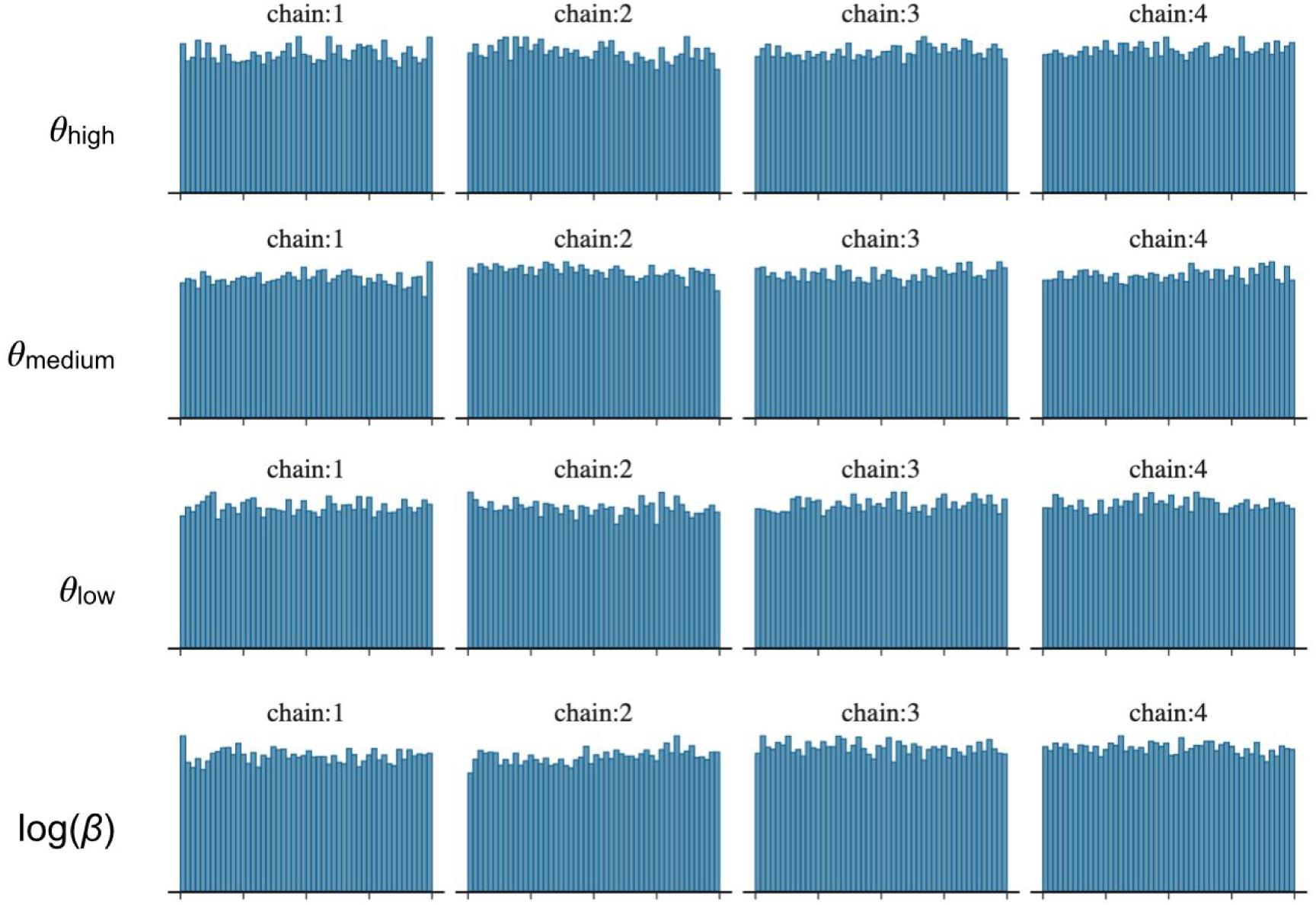
Diagnostic plots of the MCMC sampler. Shows the rank plots^12^ of the posterior samples of the four parameters used in our model. Uniform distributions indicate well-mixed MCMC chains.

